# Auto-segmentation of the parotid glands on MR images of head and neck cancer patients with deep learning strategies

**DOI:** 10.1101/2020.12.19.20248376

**Authors:** Jennifer P. Kieselmann, Clifton D. Fuller, Oliver J. Gurney-Champion, Uwe Oelfke

## Abstract

Adaptive online MRI-guided radiotherapy of head and neck cancer requires the reliable segmentation of the parotid glands as important organs at risk in clinically acceptable time frames. This can hardly be achieved by manual contouring. We therefore designed deep learning-based algorithms which automatically perform this task.

Imaging data comprised two datasets: 27 patient MR images (T1-weighted and T2-weighted) and eight healthy volunteer MR images (T2-weighted), together with manually drawn contours by an expert. We used four different convolutional neural network (CNN) designs that each processed the data differently, varying the dimensionality of the input. We assessed the segmentation accuracy calculating the Dice similarity coefficient (DSC), Hausdorff distance (HD) and mean surface distance (MSD) between manual and auto-generated contours. We benchmarked the developed methods by comparing to the inter-observer variability and to atlas-based segmentation. Additionally, we assessed the generalisability, strengths and limitations of deep learning-based compared to atlas-based methods in the independent volunteer test dataset.

With a mean DSC of 0.85*±* 0.11 and mean MSD of 1.82 *±*1.94 mm, a 2D CNN could achieve an accuracy comparable to that of an atlas-based method (DSC: 0.85 *±*0.05, MSD: 1.67 *±*1.21 mm) and the inter-observer variability (DSC: 0.84 *±*0.06, MSD: 1.50 *±*0.77 mm) but considerably faster (<1s v.s. 45 min). Adding information (adjacent slices, fully 3D or multi-modality) did not further improve the accuracy. With additional preprocessing steps, the 2D CNN was able to generalise well for the fully independent volunteer dataset (DSC: 0.79 *±*0.10, MSD: 1.72 *±*0.96 mm)

We demonstrated the enormous potential for the application of CNNs to segment the parotid glands for online MRI-guided radiotherapy. The short computation times render deep learning-based methods suitable for online treatment planning workflows.

## I. Introduction

The dramatic increase of repeated magnetic resonance imaging (MRI) in treatment planning and response assessment of radiotherapy (RT) requires the reliable segmentation of organs-at-risk (OARs) and radiation targets^1^. Manual segmentation, which is the current clinical practice, is subjective and time-consuming^2,3^. This is particularly the case for head and neck cancer (HNC) patients due to their complex anatomy which includes many OARs and several complex irradiation targets. Automating the contouring of regions of interest (ROIs) would allow to alleviate the enormous workload of manual segmentation and lead to more consistent contouring^4^.

Auto-segmentation algorithms are already implemented in most commercial treatment planning systems in the form of atlas-based methods, such as in Eclipse (Varian Medical Systems, Palo Alto, USA), Monaco (Elekta AB, Stockholm, Sweden), Pinnacle (Philips Healthcare, Best, The Netherlands) and Raystation (Raysearch Laboratories, Stockholm, Sweden). In atlas-based segmentation, the contoured anatomy of an image, also known as an atlas, is used as a source of positional and shape-based information about ROIs. Typically, multiple atlases are used, collected in a library. The contours from the library of atlases are warped to the previously unseen image using image registration methods. One then needs to select an atlas or merge multiple atlases to obtain the final contours for the unseen image. A downside of this method is the long computation time due to the multiple registrations. between the unseen image and the library of atlases.

Recent developments in the field of machine learning offer ample opportunities for automated segmentation^5^. Nowadays, the most commonly used deep learning architectures are convolutional neural networks (CNNs) with an increasing number of applications in the field of medical image segmentation. One of the most widely spread network architectures in this field is the U-Net^6^ which takes images as input and generates segmentation maps as output. Such networks are typically trained by showing pairs of images and their ground truth contours (usually generated by an expert clinician). Once the network has seen enough contours, it will be able to predict contours on new images.

Traditionally, the U-Net was developed for 2D images. However, applying deep learning to 2D slices only considers image information in 2D planes, whereas most medical imaging data are volumetric. Several studies have designed 3D CNNs^7–9^, however, feeding 3D volumes into a network consumes large computational memory and 3D convolutions are associated with a higher computational cost than their 2D counterparts. To overcome these challenges, multiple studies proposed a 2.5D approach, where images are fed into the network as adjacent slices or triplanar patches^10–13^. MRI offers the potential to acquire abundant information on the patient’s anatomy by varying image acquisition parameters. This complementary multi-modality data can be fed into the network for potentially improved performance^14,15^.

In the last couple of years, a few studies have investigated CNN-based methods for the segmentation of ROIs in HNC patients on CT images^12,16–19^ but none of them on only MR images of HNC patients. While MRI can provide a better soft-tissue contrast than CT images and therefore improve the visibility of ROIs, the resolution and contrast of MR images can substantially vary.

This study aimed to analyse whether CNN-based methods can be applied to segment the parotid glands on MR images. We hypothesise that our designed methods are at least as good as its alternatives (manual segmentation or atlas-based) but considerably faster and they reduce the workload to clinical staff. For this purpose, we used the U-Net architecture and designed and analysed the performance of 2D, 2.5D, 3D and multi-modality networks. We compared our results to a well-established multi-atlas-based algorithm and analysed the transferability of our algorithms to a fully independent test dataset.

### II. Materials and Methods

We developed all data processing tools in Python (version 3.6). Training and inference of the neural networks were performed in Tensorflow (version 1.10.0) and Keras (version 2.2.2).

### II.A. Data acquisition and preparation

Two datasets were used in this study:

- **Dataset 1:** Baseline T1- and T2-weighted MR scans of 27 HNC patients of the MD Anderson Cancer Center (Houston, Texas, USA), all with a tumour at the base of the tongue.
- **Dataset 2:** Baseline T2-weighted MR scans of 8 healthy volunteers, acquired at the Royal Marsden Hospital (RMH, London, UK).
- **Gold standard annotations:** An expert at the RMH manually contoured the left and right parotid glands using the treatment planning system Raystation (Raysearch, Stockholm, Sweden).

The patient and healthy volunteer studies were approved by the local ethics committee of the MD Anderson Cancer Center and the RMH, respectively, and all healthy and patient volunteers gave written informed consent. Figure 1 illustrates exemplary axial, sagittal and coronal views of all datasets, together with the manually segmented ROIs. Table 1 lists the relevant image acquisition parameters for each imaging modality.

**Table 1:** Imaging parameters of dataset 1 (27 patient T1w and T2w MR images) and dataset 2 (8 healthy volunteer T2w MR images) used in this study.

| parameter | dataset 1 |  | dataset 2 |
| --- | --- | --- | --- |
|  | T2w | T1w | T2w |
| vendor | GE Healthcare | GE Healthcare | Siemens |
| magnetic field strength | 3 T | 3 T | 1.5 T |
| FOV [#pixels] | 512x512 | 512x512 | 384 x 288 |
| #slices | 30 | 30 | 208 |
| voxel size [mm<sup>3</sup>] | 0.5x0.5x4 | 0.5x0.5x4 | 0.78 x 0.78 x 0.80 |
| TE [ms] | 96.72 - 107.30 | 6.54 - 7.85 | 184 |
| TR [ms] | 3198 - 4000 | 601 - 800 | 3500 |
| flip angle [°] | 90 | 90 | 120 (varying) |
| sequence type | 2D spin-echo | 2D spin-echo | 3D turbo-spin-echo |
| orientation | axial | axial | sagittal |
| #excitations (averaging) | 1 | 1 | 1.6 |

**Figure 1:**
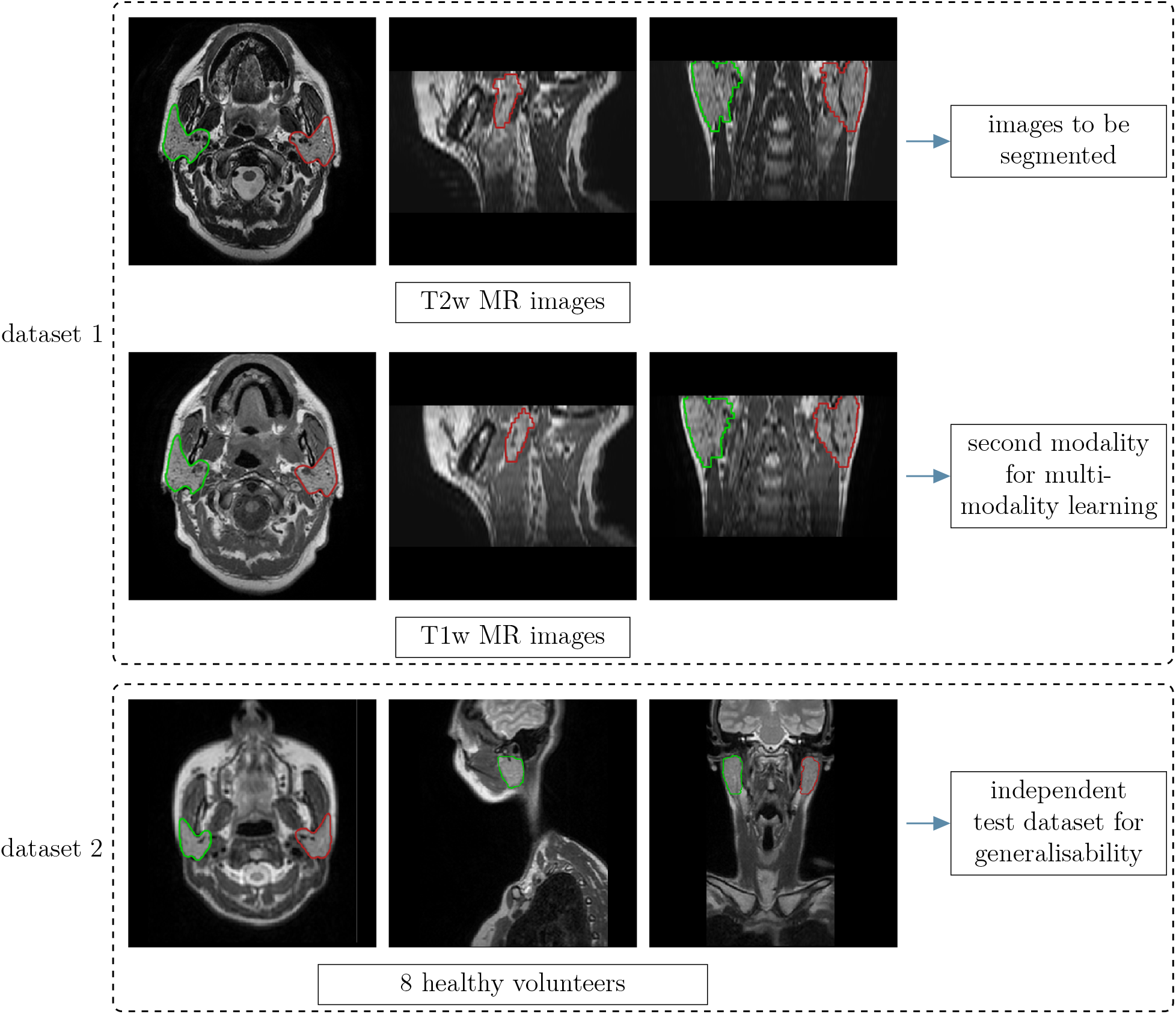
Examples of images used in this study: Axial, coronal and sagittal views of the T2-weighted and T1-weighted MR images of dataset 1 (top block), as well as the healthy volunteer and patient T2-weighted images in dataset 2 (bottom block). The coloured lines represent manually drawn contours of the left (red) and right (green) parotids.

The T1- and T2-weighted images of dataset 1 were acquired immediately after each other in the same treatment position. We further refined their alignment using a deformable image registration method of the software package NiftyReg^20,21^ (employed parameter settings: three resolution levels, control point grid size: 5×5×5mm^3^, similarity measure: normalised cross correlation, penalty terms: bending energy and Jacobian log determinant).

There were substantial differences between the two datasets, as images were acquired on scanners from different vendors and the employed sequences and image reconstruction algorithms varied. These factors contributed to differences in image quality, such as signal-to-noise ratio or spatial resolution. Such differences are representative for typical differences seen between and within clinics (e. g. due to imaging protocol or scanner upgrades). Segmentation algorithms which were developed in a particular dataset, however, may not work well in new datasets which differ significantly from the training dataset. Therefore, dataset 2 could provide valuable insights into the generalisability of developed auto-segmentation algorithms to a new dataset.

As the image intensities can vary from MR image to MR image, we standardised the contrast by performing an intensity histogram-based thresholding technique and rescaled all images to an intensity range between 0 and 255. A bottleneck in training CNNs is the GPU memory. To handle this problem and to match the images of dataset 2 to dataset 1, we resampled all images to an in-plane resolution of 1×1 mm^2^.

The images in dataset 2 covered a much larger field of view along the axial direction (head to foot). Therefore, a preprocessing step was required to avoid a failure of the image registration in the atlas-based strategy. For this purpose, we reduced the field of view in this direction to a coverage similar to dataset 1. To mimic the slight blurriness of the 3D sampling pattern in dataset 2, the images of dataset 1 (2D) were initially upsampled to a 2×2 mm^2^ in-plane resolution before being downsampled to 1×1 mm^2^ for the purpose of training the network for contouring dataset 2 (but not for the initial evaluation of contouring dataset 1).

### II.B. Neural network specifications

For the U-Net architecture^6^, we used 5 resolution levels, starting at 64 features and ending at 1024 features at the lowest resolution in the bottleneck. We employed the Adam optimiser^22^, which has proven successful in many recent applications for medical imaging.

We chose a Dice loss function^7^ as a solution to account for highly unbalanced problems where the region of interest only contributes a small percentage to the full image. With *n* denoting the number of training examples, *N*_vox_ the number of voxels, *y*^(*i*)^ the ground truth of the *i*th training example and *ŷ*^(*i*)^ the prediction of the *i*th training example, the Dice loss is defined as

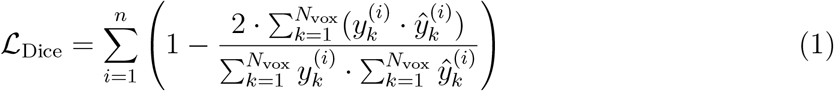

The output of the U-Net is a probability map with values between 0 and 1. We chose a threshold of 0.5 to binarise the segmentation maps. We further used a batch size of 1 for all cases.

We explored different methods, varying the dimensionality of the input, as well as adding multiple modalities. All methods are detailed in the following paragraphs and are illustrated in figures 2 and 3.

**Figure 2:**
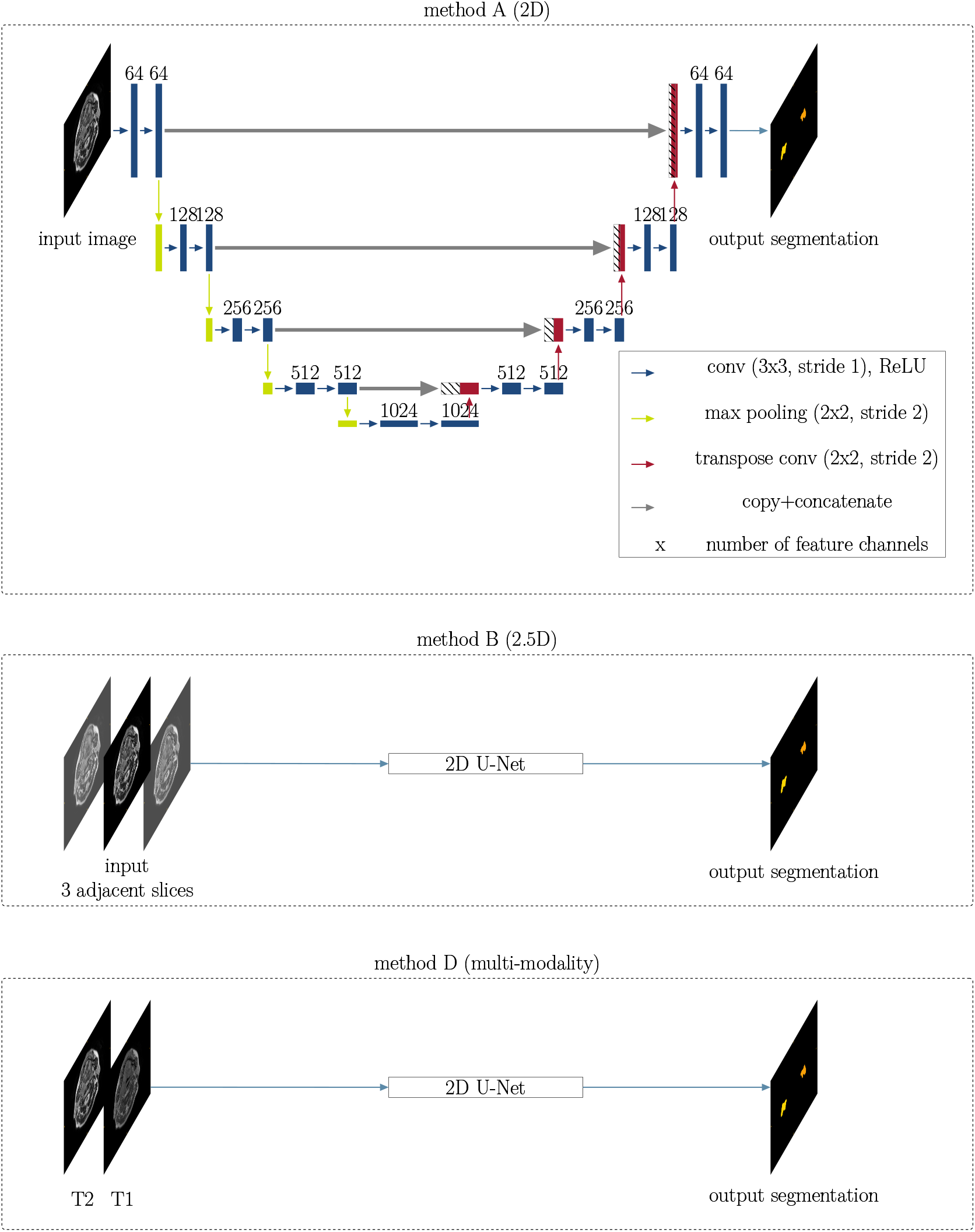
Methods using a 2D U-Net architecture: This figure illustrates methods A (2D), B (2.5D) and D (multi-modality), all using the same network architecture (2D U-Net with 5 resolution levels, starting at 64 features and ending at 1024 features at the lowest resolution in the bottleneck, comprising convolution (conv), max pooling and transpose convolution layers). Each rectangle corresponds to a feature map. The feature channels are denoted at the top of the rectangles. Striped boxes represent copied feature maps. The coloured arrows denote the different operations as indicated in the legend. The output for all three methods is a 2D segmentation map.

**Figure 3:**
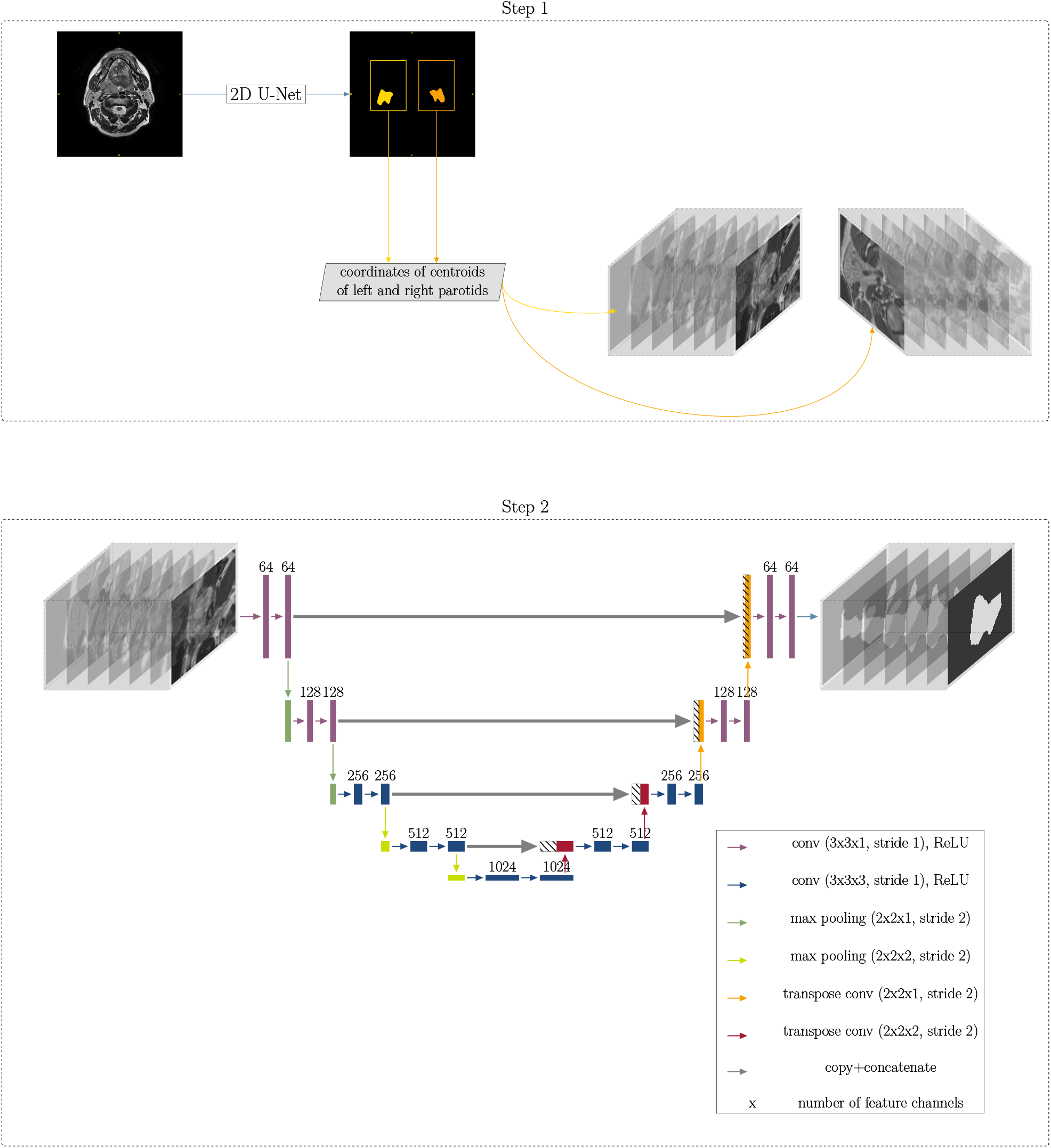
3D method: In the first step, a rough segmentation is performed (using a 2D U-Net as illustrated in figure 2) to determine the location of the left and right parotid glands. From this, 3D patches centred at the centroid coordinate of each parotid are extracted and fed into a 3D U-Net, as shown below in step 2.

### II.B.1. Method A: 2D slices

The input data to the 2D U-Net comprised the 2D axial slices of each 3D MR image in dataset 1. During training, we excluded slices which did not include any part of the parotid glands to focus the network on the relevant regions. We trained the network for 60 epochs with a learning rate of 10^*−*4^. The network is illustrated in the top part of figure 2.

### II.B.2. Method B: Adjacent slices, 2.5D

In the 2.5D network, we fed the two adjacent slices as two additional input channels into the network, as illustrated in the central part of figure 2. The network was trained for 60 epochs and a learning rate of 10^*−*4^.

### II.B.3. Method C: 3D patches with a two-step process

Due to limitations in GPU memory and to focus the network on the relevant regions of interest, we used 3D patches of 128×128×16 voxels (1×1×4 mm^3^), centred at the centre of mass of each parotid gland. To obtain the centre of mass for the testing data, we performed a rough segmentation using the 2D U-Net as a first step. Figure 3 illustrates the workflow of this method. 3D convolutions replaced the 2D convolutions of the methods described before. Due to highly anisotropic voxels, we performed the convolution in the first two levels with anisotropic convolutional kernels (3×3×1 voxels), as well as max-pooling kernels (2×2×1 voxels). In all other levels, we used isotropic kernels (convolutional: 3×3×3 voxels, max-pooling: 2×2×2 voxels).

We trained the network for 60 epochs with a learning rate of 10^*−*4^ in the first step, using downsampled images (128×128×30 voxels, 2×2×4 mm^3^ voxel size). The 3D network was trained for 40 epochs with a learning rate of 10^*−*5^.

### II.B.4. Method D: Multi-modality

To use complementary information on the regions of interest, we explored the usage of other MRI contrasts to guide the segmentation. We fed the corresponding T1- and T2-weighted images into the network as two input channels. The method is illustrated in the bottom part of figure 2. We trained the network for 60 epochs with a learning rate of 10^*−*4^.

### II.C. Atlas-based algorithm

To compare deep learning to an established algorithm, we followed an atlas-based strategy. After registering each test image to the atlas database using an affine initialisation and a deformable approach, we performed a weighted majority voting to obtain the fused segmentation. Further details are described in our previous publication^23^.

### II.D. Quantitative evaluation

For a quantitative evaluation of the auto-segmentation accuracy for dataset 1, a 9-fold crossvalidation was employed, where in each fold, three images were set aside as test images and the remaining 24 images served as training data.

### II.D.1. Computation time

The run times were determined for programme execution on a single Tesla V100 with 16 GB VRAM. We calculated the mean and average values from the individual training times of the nine folds. Inference times are stated per patient, where we calculated the average over all 27 patients and the standard deviation.

### II.D.2. Geometric evaluation

Geometric differences between the manually and the automatically derived segmentations were evaluated by calculating the 3D Dice similarity coefficient (DSC), Hausdorff distance (HD) and mean surface distance (MSD). The geometric evaluation was compared to the pairwise inter-observer variation between three observers, calculated for a subset of 12 patients of the same patient cohort^23^.

### II.E. Evaluation in independent test dataset

To assess the generalisability of deep learning-based compared to atlas-based methods, we employed the 2D method and compared it to the atlas-based method. The training data and atlas database comprised all images from dataset 1, whereas the test data comprised the images of dataset 2.

## III. Results

### III.A. Computation time

Training and inference times can be found in table 2. Methods A, B and D all had a training time of approximately 25 minutes and an inference time of less than 1 second. Method C (3D) took on average 376 minutes to train with an average inference time of 1.54 seconds. In comparison, atlas-based segmentation took approximately 45 minutes to for inference.

**Table 2:**
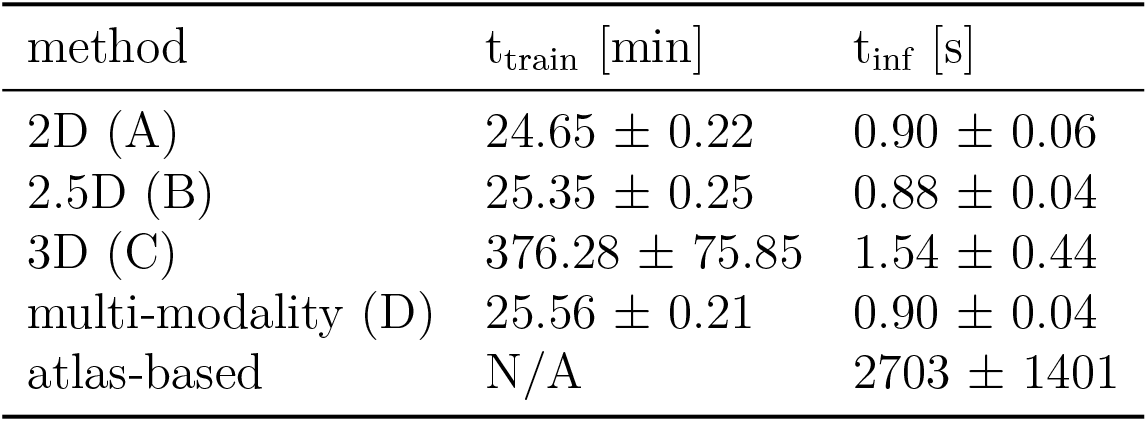
Training and inference times for the four CNN-based methods (2D, 2.5D, 3D and multi-modality), compared to the inference time for atlas-based segmentation. Training times are the average of nine folds plus/minus the standard deviation, whereas inference times are the average of 27 patients.

| method | $t_{\text{train}}$ [min] | $t_{\text{inf}}$ [s] |
| --- | --- | --- |
| 2D (A) | $24.65 \pm 0.22$ | $0.90 \pm 0.06$ |
| 2.5D (B) | $25.35 \pm 0.25$ | $0.88 \pm 0.04$ |
| 3D (C) | $376.28 \pm 75.85$ | $1.54 \pm 0.44$ |
| multi-modality (D) | $25.56 \pm 0.21$ | $0.90 \pm 0.04$ |
| atlas-based | N/A | $2703 \pm 1401$ |

### III.B. Geometric evaluation

Figure 4 shows the boxplots of all methods and ROIs employed in this study. Table 3 lists corresponding mean values and standard deviations. For comparison, we included the inter-observer variability.

**Table 3:**
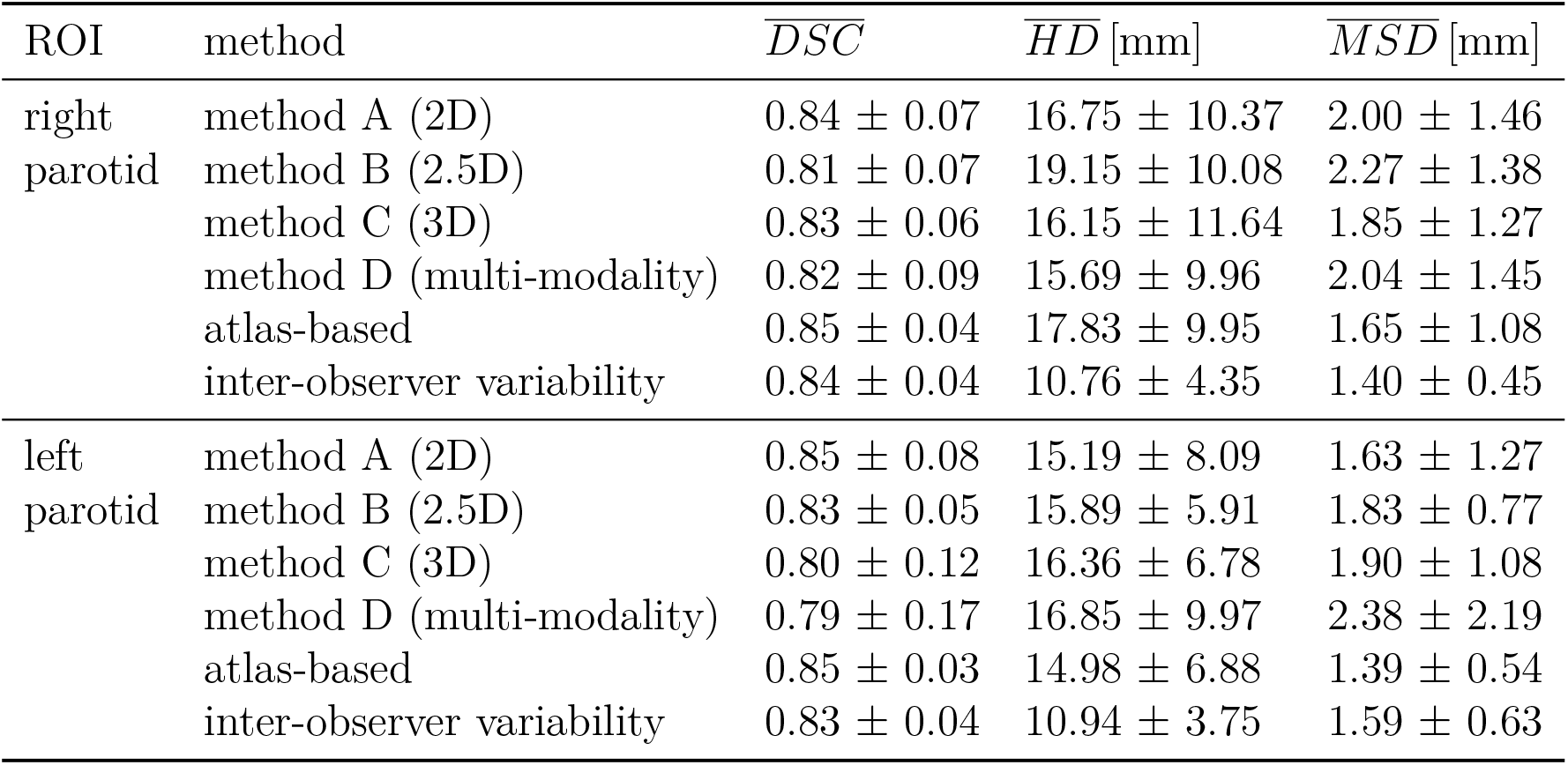
Evaluation of geometric accuracy of auto-segmenting the left and right parotid gland, comparing different methods using the U-Net (mean plus/minus standard deviation over all patients).

| ROI | method | $\overline{DSC}$ | $\overline{HD}$ [mm] | $\overline{MSD}$ [mm] |
| --- | --- | --- | --- | --- |
| right<br>parotid | method A (2D) | $0.84 \pm 0.07$ | $16.75 \pm 10.37$ | $2.00 \pm 1.46$ |
| | method B (2.5D) | $0.81 \pm 0.07$ | $19.15 \pm 10.08$ | $2.27 \pm 1.38$ |
| | method C (3D) | $0.83 \pm 0.06$ | $16.15 \pm 11.64$ | $1.85 \pm 1.27$ |
| | method D (multi-modality) | $0.82 \pm 0.09$ | $15.69 \pm 9.96$ | $2.04 \pm 1.45$ |
| | atlas-based | $0.85 \pm 0.04$ | $17.83 \pm 9.95$ | $1.65 \pm 1.08$ |
| | inter-observer variability | $0.84 \pm 0.04$ | $10.76 \pm 4.35$ | $1.40 \pm 0.45$ |
| left<br>parotid | method A (2D) | $0.85 \pm 0.08$ | $15.19 \pm 8.09$ | $1.63 \pm 1.27$ |
| | method B (2.5D) | $0.83 \pm 0.05$ | $15.89 \pm 5.91$ | $1.83 \pm 0.77$ |
| | method C (3D) | $0.80 \pm 0.12$ | $16.36 \pm 6.78$ | $1.90 \pm 1.08$ |
| | method D (multi-modality) | $0.79 \pm 0.17$ | $16.85 \pm 9.97$ | $2.38 \pm 2.19$ |
| | atlas-based | $0.85 \pm 0.03$ | $14.98 \pm 6.88$ | $1.39 \pm 0.54$ |
| | inter-observer variability | $0.83 \pm 0.04$ | $10.94 \pm 3.75$ | $1.59 \pm 0.63$ |

**Figure 4:**
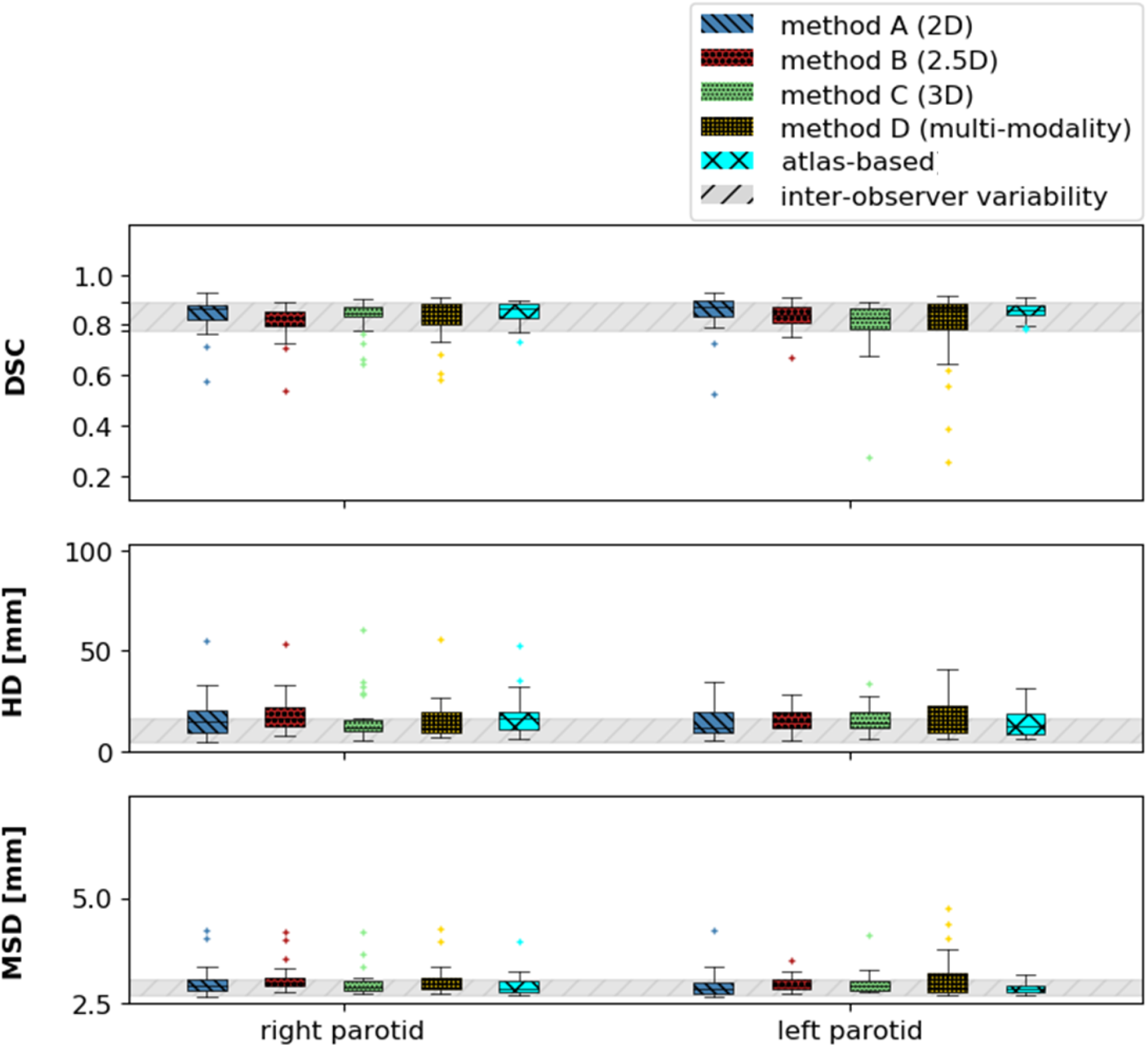
Comparison of different CNN-based methods: Boxplots of, from top from bottom, the DSC, the MSD and the HD for both parotid glands (x-axis) and all automated segmentation methods (A in blue, B in red, C in green, D in yellow). We also included the inter-observer variability (grey).

With a mean DSC larger than 0.8 and a mean MSD smaller than 2.5 mm, all methods achieved a geometric accuracy which was in the same ballpark as the inter-observer variability. There were no significant differences in the average performance of any of the methods. While the 3D method tended to have a smaller variance of accuracies and less severe misclassifications, the multi-modality method added some uncertainty in comparison to the plain 2D method. Adding adjacent slices did not improve the segmentation accuracy.

### III.C. Typical segmentation examples

Figure 5 provides four selected examples of the auto-generated contours overlayed with the manual contours, comparing all four methods (2D, 2.5D, 3D and multi-modality). The rows represent the four methods, whereas the columns show four patients. The contours generally followed the manual contours well. Patient 1 and 2 are typical examples. The auto-segmentation of patient 3 (third column) illustrates a case where methods C (3D) and D (multi-modality) did not perform well: for method C, most of the left parotid is left out, being entirely missed out for method D. Patient 4 (last column) is an example where it was difficult to tell whether the auto-generated or manual contours were the more accurate ones: the part included additionally for the right parotid in the auto-generated contours in comparison to the manual ones might as well be part of the parotid gland which might have been missed out in the manual procedure.

**Figure 5:**
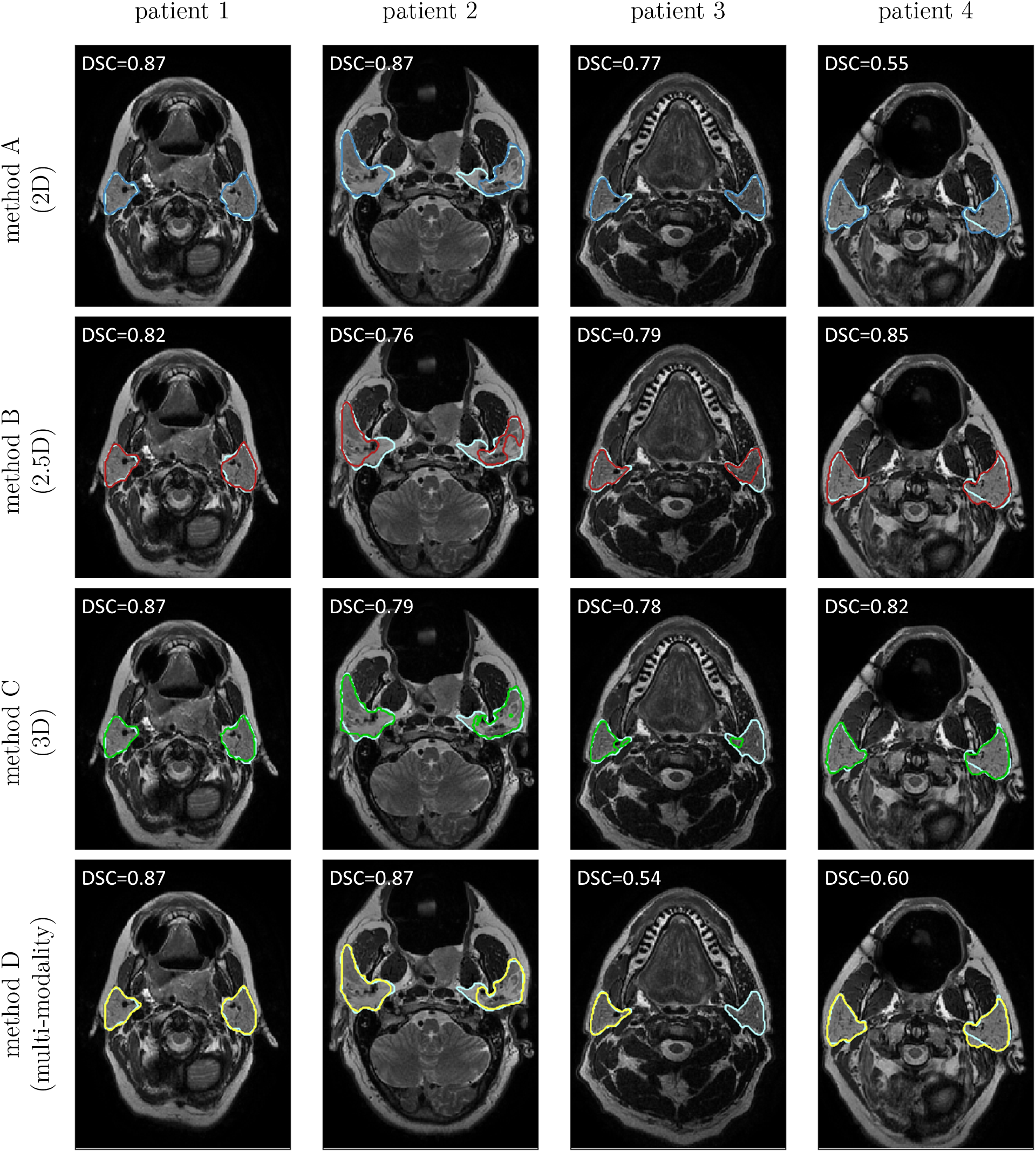
Deep learning-based segmentation: Each column displays a typical example comparing the manually segmented parotids (light blue) to method A (2D, dark blue, first row), method B (2.5D, red, second row), method C (3D, green, third row) and method D (multimodality, yellow, fourth row), respectively. Each example originates from a different patient. The figure illustrates 2D slices, whereas the indicated DSC was calculated for the full 3D volume.

Figure 6 illustrates typical examples where parotid glands were infiltrated by involved lymph nodes. In general (e. g. columns 1-3), the deep learning-based methods seemed to be better at detecting boundaries of parotid glands that were infiltrated by involved lymph nodes or primary tumours and did not include them within the segmented ROI. The last column in figure 6 represents a rare example where the atlas-based outperformed the deep learning-based method. In such a situation, the lymph nodes appeared to have a similar texture compared to the parotid gland itself, which was likely the reason that the network annotated this part as parotid gland.

**Figure 6:**
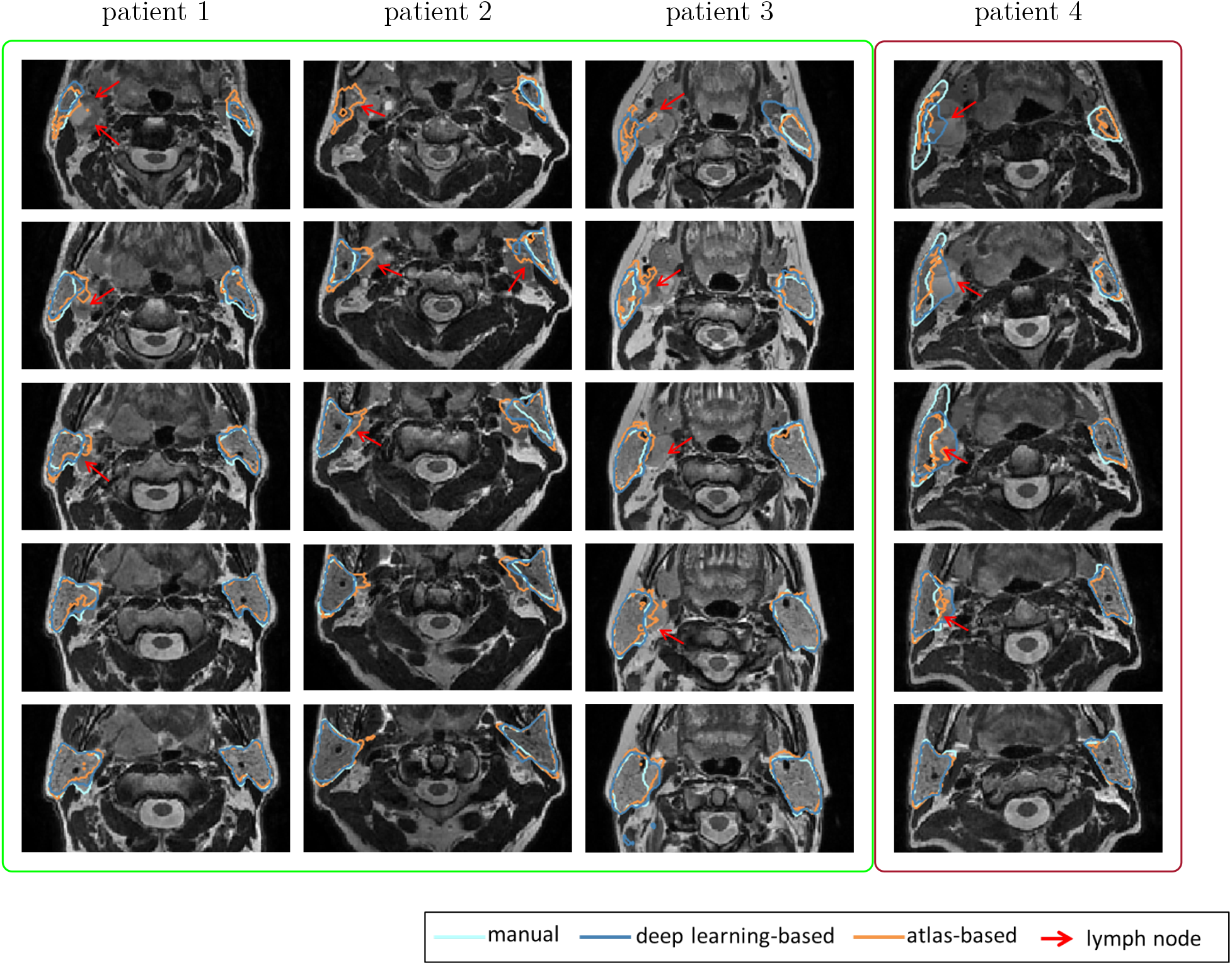
Segmentation examples: This figure displays four cases where the parotid glands were infiltrated by involved lymph nodes. Each column shows an example case with consecutive axial slices shown in each row. The different colours indicate different methods (dark blue: 2D deep learning method, orange: atlas-based method, light blue: manual gold standard). The red arrows point towards involved lymph nodes. The first three columns highlight examples where deep learning outperformed an atlas-based method (green box) whereas the last column provides a less common counter-example (red box).

### III.D. Evaluation in independent test dataset

Figure 7 illustrates typical example cases comparing the atlas-based method to the deep learning-based method for the independent test dataset (dataset 2). The deep learning-based contours appeared less fuzzy at the edges compared to the atlas-based contours. Most of the atlas-based generated contours followed the boundaries of the parotid glands closely.

**Figure 7:**
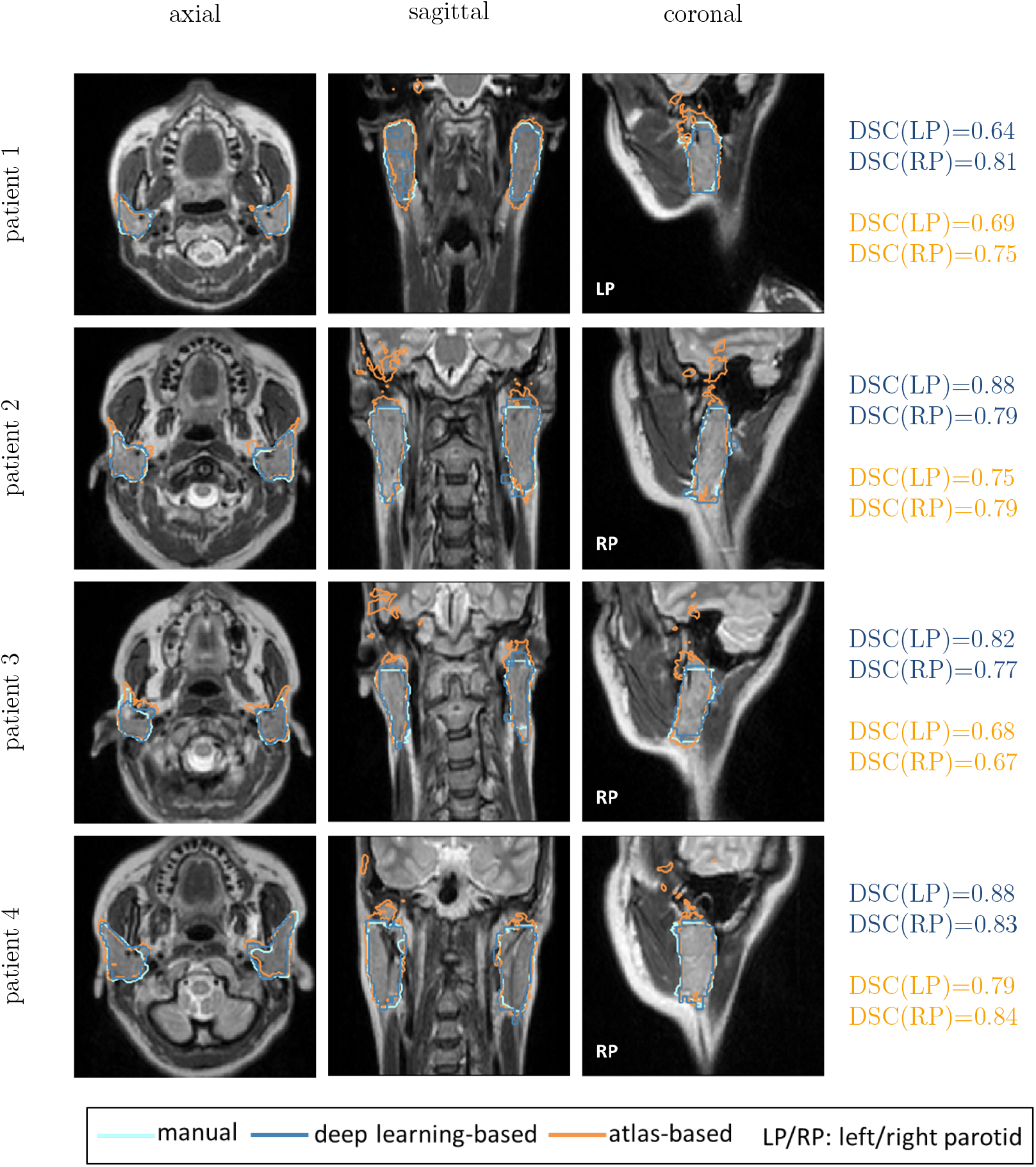
Segmentation examples: this figure provides 4 example cases for atlas- and deep learning-based auto-segmented left (LP) and right parotid (RP) glands (orange: atlas-based, dark blue: deep learning-based, light blue: manual gold standard). The rows each show a typical example, whereas the columns illustrate the axial, sagittal and coronal cross-sections, respectively.

Table 4 lists values for the geometric measures of deep learning compared to atlas-based segmentation results. The deep learning-based method achieved a higher accuracy (DSC: 0.79*±*0.10, HD: 11.91*±*8.41 mm, MSD: 1.72*±*0.96 mm) compared to the atlas-based method (DSC: 0.70*±*0.16, HD: 43.98*±*21.16 mm, MSD: 5.76*±*6.36 mm).

**Table 4:** Evaluation of geometric accuracy of auto-segmenting the left and right parotid gland for the independent test dataset, comparing a 2D deep learning method with atlas-based (mean plus/minus standard deviation of all patients).

| ROI | method | $\overline{DSC}$ | $\overline{HD}$ [mm] | $\overline{MSD}$ [mm] |
| --- | --- | --- | --- | --- |
| right | deep learning (2D) | $0.78 \pm 0.06$ | $12.61 \pm 7.75$ | $1.79 \pm 0.75$ |
| parotid | atlas-based | $0.72 \pm 0.12$ | $44.48 \pm 16.55$ | $5.14 \pm 5.24$ |
| left | deep learning (2D) | $0.79 \pm 0.08$ | $11.20 \pm 3.28$ | $1.64 \pm 0.60$ |
| parotid | atlas-based | $0.68 \pm 0.10$ | $43.48 \pm 13.19$ | $6.38 \pm 3.60$ |

## IV. Discussion

We investigated the application of CNN-based methods to the segmentation of the parotid glands and benchmarked them with the inter-observer variability, as well as an atlas-based method. To our knowledge, this was the first study to demonstrate that auto-contouring of the parotid glands on MR images using CNN-based methods can achieve an accuracy comparable to the inter-observer variability. We found that 2D CNNs were able to perform as well as the current benchmarks but considerably faster (<1s v.s. 45 min). This renders deep learning-based methods feasible in an online adaptive clinical workflow, such as envisioned with the MR-Linac^24–27^. We also found that adding neighbouring slices (2.5D/3D) or multiple modalities did not further improve the performance of CNNs for this purpose.

We compared four different methods: 2D (method A), 2.5D (method B), 3D (method C) and multi-modality (method D). All methods achieved an accuracy that was of the same size as the inter-observer variability with a DSC larger than 0.8 and an MSD smaller than 2.5 mm. There were no clear benefits of using one over the other for the data in this study. Since all methods achieved an accuracy that was already in the same range as the inter-observer variability, it was challenging to detect differences in the methods.

Combining multiple modalities could provide the network with complementary information about tissues. We, therefore, explored how the combination of T1-weighted and T2-weighted contrast could improve auto-segmentation in method D. This approach did not increase the accuracy but instead led to more misclassified voxels. This may be attributed to the finding that the T2-weighted and corresponding T1-weighted images may not have been correctly aligned, due to motion, for example, despite the images been taken subsequently, within the same treatment position, and an additional registration between the images, and therefore led to inconsistent information for the CNN.

Adding information of neighbouring slices also did not improve the performance. The 3D method decreased the general spread of values, but the overall accuracy did not improve in comparison to the 2D method. Due to memory-related restrictions of the hardware, we followed a two-step approach by first finding an approximated bounding box close to the parotids and then using a 3D patch with the centroid at this bounding box. This requires the first step to being sufficiently accurate as otherwise the parotid might be missed in the extracted 3D patch. However, we can confirm that the 2D approach of this study always gave the correct location to find the centre of mass for the 3D method.

Table 5 lists mean reported geometric measures for a comparison of our results to published studies on CNN-based methods applied to the segmentation of the parotid glands. All of the reported studies used CT images (with one study adding MR images in a multi-modality method). In particular, the 2D method ranked amongst the top of the published studies with only the study by Chan et al.^18^ outperforming it. The fact that our method performed well may be related to the fact that the parotid glands are better visible on MR images and all included studies used CT images. The training data of published studies were of similar size as ours except for two studies which had larger datasets^18,19^. We expect that with more training data, one can reduce the number of misclassified voxels and hence improve performance. This is reflected in the CT studies performing better with larger datasets in table 5. This also explains why the CT study by Chan et al.^18^, which included 200 patients, performed slightly better than our MRI approach with only 27 patients.

**Table 5:** Comparison of geometric evaluation for the methods developed in this study to published studies

| ROI | DSC | MSD[mm] | mod | # | study |
| --- | --- | --- | --- | --- | --- |
| right | $0.84 \pm 0.07$ | $2.00 \pm 1.46$ | MR | 27 | method A (2D) |
| parotid | $0.81 \pm 0.07$ | $2.27 \pm 1.38$ | MR | 27 | method B (2.5D) |
| | $0.83 \pm 0.06$ | $1.85 \pm 1.27$ | MR | 27 | method C (3D) |
| | $0.82 \pm 0.09$ | $2.04 \pm 1.45$ | MR | 27 | method D (multi) |
| | $0.85 \pm 0.04$ | $1.65 \pm 1.08$ | MR | 27 | atlas-based |
| | $0.84 \pm 0.02$ | $1.12 \pm 0.56$ | CT | 32 | Tong et al.<sup>16</sup> |
|  | 0.79 | 1.57 | CT+MR | 43 | Mocnik et al.<sup>17</sup> |
| | $0.78 \pm 0.05$ | - | CT | 50 | Ibragimov et al.<sup>12</sup> |
| | $0.86 \pm 0.05$ | - | CT | 200 | Chan et al.<sup>18</sup> |
| | $0.83 \pm 0.02$ | - | CT | 157 | van Rooij et al.<sup>19</sup> |
| left | $0.85 \pm 0.08$ | $1.63 \pm 1.27$ | MR | 27 | method A (2D) |
| parotid | $0.83 \pm 0.05$ | $1.83 \pm 0.77$ | MR | 27 | method B (2.5D) |
| | $0.80 \pm 0.12$ | $1.90 \pm 1.08$ | MR | 27 | method C (3D) |
| | $0.79 \pm 0.17$ | $2.38 \pm 2.19$ | MR | 27 | method D (multi) |
| | $0.83 \pm 0.06$ | $1.65 \pm 1.57$ | MR | 27 | atlas-based |
| | $0.84 \pm 0.03$ | $0.96 \pm 0.34$ | CT | 32 | Tong et al.<sup>16</sup> |
|  | 0.79 | 1.57 | CT+MR | 43 | Mocnik et al.<sup>17</sup> |
| | $0.77 \pm 0.06$ | - | CT | 50 | Ibragimov et al.<sup>12</sup> |
| | $0.85 \pm 0.03$ | - | CT | 200 | Chan et al.<sup>18</sup> |
| | $0.83 \pm 0.03$ | - | CT | 157 | van Rooij et al.<sup>19</sup> |

As shown in figure 6, the atlas-based method often performed worse compared to the deep-learning method when anatomy was disrupted, for instance, by involved lymph nodes. Since the shape and location of tumours and involved lymph nodes varies widely among patients, it is difficult for an atlas-based method to recognise infiltrated normal structures. Deep-learning strategies, on the other hand, are recognising local shapes and edges and are, therefore, less affected by global geometrical changes, as shown by typical examples in figure 7. We, therefore, believe that deep learning-based approaches can be expected to outperform atlas-based approaches when applied to, and in the presence of, irregular structures such as tumours.

Performances of networks can depend on how they are trained, as indicated by the hyperparameters. During the selection of suitable hyperparameters, we explored the application of data augmentation (random translations, rotations, mirroring, random cropping) and drop-out at different places within the network, but did not see any improvement in the performance of the network. We also tried an alternative loss function based on the weighted cross-entropy, which did not improve the network’s performance. As the data augmentation and the alternative loss function did not improve the networks performance, we decided to take the most basic approach with the minimum number of tweakable hyperparameters. These exploratory approaches were only done with only a subset (1/9) of the data for testing to minimise leakage and over-optimising hyperparameters to our data.

In clinical practice, images can vary in their signal-to-noise ratio, resolution or image quality due to protocol or scanner upgrades within a clinic or differences in protocols or scanners between clinics. For this purpose, we employed an independent test dataset in this study. Although this dataset also comprised T2-weighted MR images, it was substantially different from the training dataset. Images were obtained with different setup protocols in different hospitals. As the dataset was acquired at different field strengths and as a 3D acquisition (considerably longer TEs and variable flip angles) instead of 2D, the contrast was different. Furthermore, it was obtained at a different resolution, with further blurring due to the long scan times and signal intensity variations throughout k-space associated with the 3D approach. Therefore, it is very promising that, with some minor obvious modifications of the training data (resampling, blurring) the network performed well on this independent testing data. However, without these adjustments, the network failed in the new patients (results not shown). That said, the atlas-based approach also required adjustments of the data (smaller FOVs). For more complicated cases, where the contrast could have changed substantially, a promising solution could be the cross-modality learning approach^28^ where one could transfer the expertise gained in the contouring of one type of sequence to any desired sequence through synthetic image generation.

The deep learning-based method outperformed the atlas-based method in the independent test dataset and could achieve an accuracy which was close to the inter-observer variability of the patient dataset.

## IV.A. Limitations of this study

In this study, we focused on the segmentation of the parotid glands. While the segmentation of these OARs is crucial, the methodology from this study is not limited to this specific organ and we expect that one can transfer the method to the segmentation of other ROIs. Out of the many OARs relevant for the treatment planning of RT in HNC, the parotid glands are the most challenging ones to contour due to variations in shape and location. Furthermore, including multiple OARs as different labels in one network could improve the geometric accuracy as more information would be available to the CNN and the amount of misclassified voxels may be reduced.

## V. Conclusion

This study demonstrated that CNNs are able to segment the parotid glands for RT treatment planning purposes with an accuracy comparable to labour intense manual contouring. Furthermore, CNNs performed equally well as a conventional atlas-based method, however, the atlas-based method took substantially longer (<1s compared to 45 mins). These short computation times render deep learning-based methods suitable for online adaptation work-flows.

## Data Availability

The data that support the findings of this study are available from the MD Anderson Cancer Center but restrictions apply to the availability of these data, which were used under license for the current study, and so are not yet publicly available.

## VI. Acknowledgements

We would like to thank Brian Hin for his help in manually contouring all the images. We are grateful to Mona Kamal Jamaa and Yao Ding for helping with the data acquisition and export at MD Anderson, as well as to the radiographers at the RMH for their support in acquiring images of the healthy volunteers. We would like to acknowledge NVIDIA for their support of this work through donation of a Graphics Card in the GPU Grant. This work was supported by the Oracle Cancer Trust, as well as the Cancer Research UK Programme (grants C7224/A23275, and C33589/ A19727). The ICR/RMH is part of the Elekta MR-Linac Research consortium. This report is independent research funded by the National Institute for Health Research. The views expressed in this publication are those of the authors and not necessarily those of the NHS, the National Institute for Health Research or the Department of Health. We would further like to acknowledge CRUK and EPSRC support to the Cancer Imaging Centre at ICR and RMH in association with MRC and Department of Health C1060/A10334, C1060/A16464 and NHS funding to the National Institute for Health Research (NIHR) Biomedical Research Centre and the Clinical Research Facility in Imaging. CDF would like to acknowledge funding from NIH grants with numbers R25EB025787 and R01DE028290.

